# Economic Burden of Typhoid among Hospitalized Children in Kenya

**DOI:** 10.64898/2025.12.10.25341952

**Authors:** Prashant Mandaliya, Stacey Orangi, Isaac Waluke, Franklin Okech, Felix Masiye, Obinna Onwujekwe, Lumbwe Chola, Edwine Barasa

**Affiliations:** Health Economics Research Unit, KEMRI Wellcome Trust Research Programme, Nairobi, Kenya; Health Services Unit, KEMRI-Wellcome Trust Research Programme, Nairobi County, Kenya; Department of Economics, School of Humanities and Social Sciences, University of Zambia, Zambia; University of Nigeria, Nsukka, Nigeria; Bergen Centre for Ethics and Priority Setting, University of Bergen, Norway; Centre for Global Health and Tropical Medicine, Nuffield Department of Medicine, University of Oxford, Roosevelt Drive, Oxford OX3 7LG, UK

## Abstract

Typhoid remains endemic in many low and middle-income countries, with the highest incidence reported in children under 15 years. No published study has estimated the cost of hospitalized paediatric typhoid cases in Kenya. This analysis estimated the direct medical, direct non-medical, and indirect costs from the healthcare provider and societal perspectives and assessed the proportion of households that would experience catastrophic health expenditure (CHE) due to the disease.

Patient-level data on hospitalized children with typhoid were drawn from 16 public hospitals that are part of the Clinical Information Network (CIN) in Kenya. A cost-of-illness analysis was conducted. Resource quantities were extracted from CIN data, and unit costs were obtained from price lists, surveys and expert interviews. All costs were converted to 2025 KES and USD. A simulation-based catastrophic health expenditure analysis was conducted. One-way sensitivity analysis varied hospital bed, staff, non-medical and indirect costs.

Median cost per unique admission was USD 79.68 from the provider and USD 96.95 from the societal perspective. Staff costs accounted for up to 55% of societal costs, while hospital-bed costs accounted for up to 15%. Productivity loss represented 12% of societal costs. In the sensitivity analysis, adjusting staff time increased the median cost by 89% for the provider perspective and 74% for the societal perspective. Conversely, varying median daily bed charges between the 25th and 75th percentiles altered the cost of treatment by -9% to 5% for the provider and -5% to 5% for the societal perspectives. CHE incidence rose as the share paid out-of-pocket increased and house wealth declined, mainly impacting rural and poorer households.

Typhoid imposes a substantial economic burden among hospitalized children in Kenya, driven mainly by staff and hospital-bed costs. The findings of this study can guide policymakers in reducing strain on health services and the financial burden on families.

**What is already known on this topic:**

- Typhoid is endemic in Kenya, and although its epidemiology is well described, the economic burden has not been quantified.
- The gap limits the evidence available to guide financing decisions and investments in prevention strategies.

**What this study adds:**

- This study provides the first estimate of the economic burden of hospitalised paediatric typhoid in Kenya, identifies key cost drivers, and notable cost differences across geographic settings and patient groups.
- It also highlights the financial strain that typhoid can place on affected households.

**How this study might affect research, practice or policy:**

- The study provides researchers with a methodology for assessing catastrophic health expenditure when household level data is not available.
- The outputs of the study can be used to evaluate the value of vaccines for typhoid in similar contexts.
- These findings offer policymakers, local evidence to guide decisions for insurance and health financing, especially in settings and patient groups where the financial burden is greatest.

## Introduction

Typhoid fever is a systemic infection caused by *Salmonella enterica* serovar Typhi (S. Typhi) and remains endemic in many low- and middle-income countries (LMICs) [1]. Treatment initiated with appropriate antibiotics has been shown to be effective, however with the rise of antimicrobial resistance (AMR), cheaper first line treatment options are becoming less effective [1]. A study by Breiman et al. highlighted this by reporting that nearly 75% of S. Typhi isolates were resistant to multiple antimicrobials in Kenya [2]. Untreated cases can progress to more severe complications such as intestinal haemorrhage or perforation [3].

In 2021, there were about 9.3 million cases and 107,000 deaths reported globally for typhoid [4]. When incidence of the disease exceeds 100 cases per 100,000 person-years in a country, it is termed as highly endemic [5]. This level of endemicity is seen across multiple countries in Asia and sub-Saharan Africa, with Kenya identified as one of the few high incidence countries outside South Asia [6]. The estimated incidence in Kenya was estimated to be 217 cases per 100,000 persons-years [4], rising to 843 cases per 100,000 person-years in some informal settlements [2]. Urban areas were also seen to have a 10-fold higher incidence when compared to rural settings in the country [2]. The disease is concentrated in children under 15 years, who accounted for 62% of total cases and 66% of total deaths, globally [7]. A similar trend was observed in Kenya as reported by Simiyu et al., finding the highest incidence among children aged 2-4 years [7].

Quantifying the economic burden of a disease through a cost-of-illness (COI) study provides evidence to inform resource allocation and inputs for cost-effectiveness analysis of vaccines [8]. These analyses systematically identifies different costs based on the chosen perspective, aiming to determine the total cost associated with treating the disease [8]. These costs are placed in three categories: direct medical costs (healthcare expenditures for diagnosis, treatment, and rehabilitation), direct non-medical costs (non-healthcare costs for transportation, food, lodging, and caretaker fees), and indirect costs (productivity losses due to illness, caregiving, or premature death) [8]. A recent review highlighted the variability in the cost of treating typhoid across countries as well as within countries [9]. Therefore, context-specific estimates are essential, as they provide policymakers with local evidence on the economic burden of the disease in the country.

The financial impact of a disease on households can also be severe, leading to catastrophic health expenditure (CHE). CHE is defined as the costs related to healthcare that exceeds a household’s capacity to pay (CTP) [10,11], which is typically defined as 40% of the annual non-food household expenditure (NFHE) [12]. Evidence on CHE can help policymakers understand the level of OOP spending that households of different wealth levels can sustain without facing catastrophic costs. A study from Malawi indicated that 44% of households faced CHE when a household member was treated for typhoid [13], while a multi-country analysis by Poulos et al. [14] showed that treatment costs can account for nearly 15% of a household’s yearly income.

Kenya provides a critical setting for this analysis, given the high incidence of typhoid in children [4]. While the epidemiological burden of paediatric typhoid in Kenya is well documented [2,15], there is no published literature on it’s economic burden. To the best of our knowledge, this study provides the first estimate of the economic burden of typhoid fever among hospitalized paediatric cases in Kenya. The study also estimates the proportion of households likely to experience CHE under different scenarios. Policymakers can use the evidence generated from this study to better understand the economic burden of typhoid and address key cost drivers, while researchers could use the findings to evaluate the cost-effectiveness of typhoid conjugate vaccine in similar contexts.

## Methodology

### Study setting

Kenya is a LMIC that is governed through a devolved system comprising of a national government and 47 semi-autonomous counties. Health services are delivered through both the public and private sectors. Responsibilities within the public health sector are shared between the two levels of government, with counties overseeing 70% of health functions and the national government managing the remaining 30% [16]. Services provided by county health facilities are primarily funded through county health budgets, with additional funding from the social health authority (public insurer) and out-of-pocket payments from patients [17]. The Ministry of Health receives allocation through the national budget to manage national-level health functions. The national government provides funding to the State Department of Medical Services and State Department of Public Health and Professional Standards to oversee national health functions [18].

### Study design and study population

A retrospective COI study design was used to estimate the cost of treatment for a unique hospitalised typhoid case among children under 15 years. CHE incidence was assessed using a simulation-based methodology. Primary patient-level data were obtained from the Clinical Information Network (CIN), a hospital-based platform that collects routine admission, care and discharge data from neonatal and paediatric wards in 24 public hospitals across 19 (out of 47) counties in Kenya [19]. It provided a representative sample of paediatric admissions in the country.

### Costing approach, perspective, and time horizon

A bottom-up micro-ingredient costing approach was employed, where individual resources used for the treatment of typhoid were identified, measured, and valued to determine the cost of treatment [20]. The analysis was conducted from the healthcare provider and societal perspectives, with a time horizon of the patient’s hospitalization period.

### Cost components

Full economic costs were estimated by including the monetary value of all inputs, whether financial payments were involved or not. Costs were classified as direct medical costs (light grey), direct non-medical costs (medium grey), and indirect costs (dark grey) as shown in Figure 1. From the healthcare providers’ perspective, only direct medical costs were included, while the societal perspective was broader and included direct medical, direct non-medical, and indirect costs. Direct medical costs comprised of hospital bed, laboratory and radiology, medication, nutrition, blood transfusion, oxygen supplementation, and staff costs. Direct non-medical costs accounted for transport, food, commercial diapers, childcare for other dependents, companion costs and other miscellaneous costs, whereas indirect costs captured the time lost by caregivers during the child’s hospitalization period. From the healthcare providers’ perspective, only direct medical costs were included, while the societal perspective was broader and included direct medical, direct non-medical, and indirect costs.

**Figure 1:**
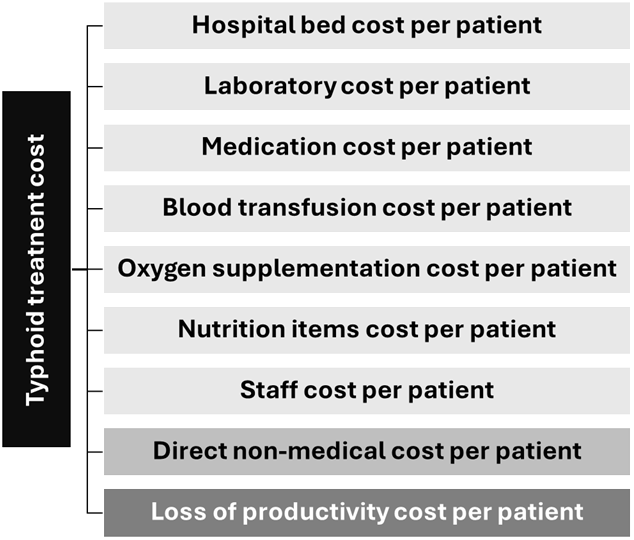
Cost categories for the treatment of typhoid

### Data collection

Data on patients diagnosed with typhoid were collected by the CIN team. This data was then extracted for patients admitted from the CIN database (hereafter referred to as CIN data), which ranged from 2014 to 2024.

The extracted data was cleaned and missing values imputed using STATA v17 [21]. The cleaning process involved removal of duplicates, standardization of diagnostic codes, and resolving treatment inconsistencies. Missing data were imputed using the mean, mode, or literature-based assumptions. These assumptions were first validated through consultations with specialists, medical officers, pharmacists, and nurses to ensure their suitability for the Kenyan context.

### Measuring and valuing resources

Resource utilization per patient was quantified primarily from the CIN data and supplemented by expert opinions. Unit costs were obtained from secondary data, market surveys, and price lists.

All costs were first converted to Kenya shillings (KES) based on the average annual exchange rate and then inflated to 2025 KES using GDP deflators for Kenya [22,23]. For the years 2024 and 2025 an annual inflation rate of 5.3% was used as GDP deflator values were missing [24]. All cost estimates reported in 2025 KES were subsequently converted to USD using an exchange rate of KES 129.28 per USD [25].

The total cost for each item (*i)* was calculated by multiplying the unit cost of the item by the total quantity consumed:

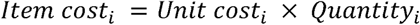

Items belonging to the same cost category (k) were then summed to obtain the total cost for each cost category:

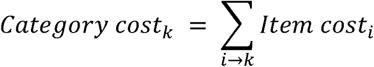

The sub-sections below outline how each cost category was measured and valued:

#### i) Hospital bed costs

The duration of hospital stay for each patient was obtained from the CIN data set using the *date of admission* and *date of discharge* variables. Daily bed costs were obtained from 14 hospitals within CIN (see Appendix 1). The median value was used as a representative value for hospital bed costs for all patients.

#### ii) Pharmaceutical, non-pharmaceutical and nutrition item costs

The quantity of pharmaceuticals consumed was determined by reviewing the variables that described the prescription, which included the drug name, route, strength, frequency, and duration. Total quantity of pharmaceuticals consumed per patient was calculated using the equation below, with the days of therapy defined as either the number of days prescribed or duration of hospitalization (whichever was shorter):

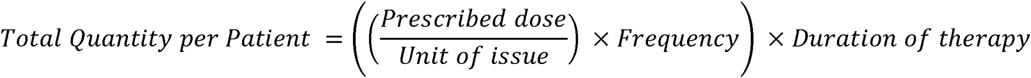

For non-pharmaceutical and nutritional items, the quantities were determined based on the frequency with which they were prescribed.

The process of obtaining prices of pharmaceuticals, non-pharmaceuticals, and nutritional items was based on the process followed at public hospitals in Kenya. The primary source was the Kenya Medical Supplies Authority (KEMSA) pricelist, which was chosen given KEMSA’s role in public procurement for public health facilities in Kenya [26]. For items not available at KEMSA, the Mission for Essential Drugs and

Supplies pricelist [27], and costs for any remaining items were obtained through a market survey of local suppliers in Nairobi, using convenience sampling [28].

#### iii) Laboratory, radiology, blood transfusions and oxygen costs

The quantities of laboratory tests, radiological procedures, blood transfusions, and oxygen supplementation were based on the number of times they were requested, for each patient.

Unit costs for laboratory and radiology services were sourced from a 2023 cost survey conducted across seven public hospitals in Kenya [29], with median values used to represent the unit cost. For tuberculosis-related tests, which are provided for free in public facilities, costs from the private sector were obtained [30]. These costs are detailed in Appendix 2.

For the unit cost of blood transfusion, a hospital survey was conducted across three public hospitals, and the reported median cost of USD 7.74 was used (see Appendix 3). The cost of oxygen supplementation was obtained from Kenya-specific literature [31].

#### iv) Staff costs

Staff time estimates were obtained from expert opinions. Purposive sampling was employed until saturation was reached [32]. Public sector health workers (n=15), which included specialists, medical officers, clinical officers, nurses, and nutritionists estimated the time health workers from their cadre would spend on a typhoid patient within a 24-hour period to cover routine care tasks such as assessment, monitoring, prescribing, administering medications, counselling, and documentation. The average reported times across the different cadres were used to quantify the staff time (see Appendix 4).

Unit costs were calculated by converting monthly salaries into per minute values, based on the assumption that a healthcare worker would work 40 hours per week in accordance with the Kenyan human resource policies [33]. Monthly salaries were sourced from a study by Barasa et al. [31], which reported official Kenyan public sector salaries for each cadre.

#### v) Direct non-medical costs

Direct non-medical costs were sourced from Kenya specific literature [34]. Per-day direct non-medical costs for urban and rural counties were calculated by first determining the weighted average cost and length of stay (LOS) for the counties based on their geographic setting. The weighted average cost was divided by the weighted average LOS to determine the cost per day for both rural and urban settings. The resulting values were adjusted to the 2025 KES value and then converted to USD (Urban: USD 4.59; Rural: USD 2.18). The duration of hospitalization served as a proxy for the quantity.

#### vi) Loss of productivity costs

Indirect costs were estimated using the human capital approach [35]. The daily wage rates for general labourers in Kenya, disaggregated by county, were used for the unit cost of productivity loss [36]. The quantity was equal to the duration of hospitalization for each patient.

### Cost function

The total cost of illness was calculated by summing the cost categories for each patient, based on the perspective employed:

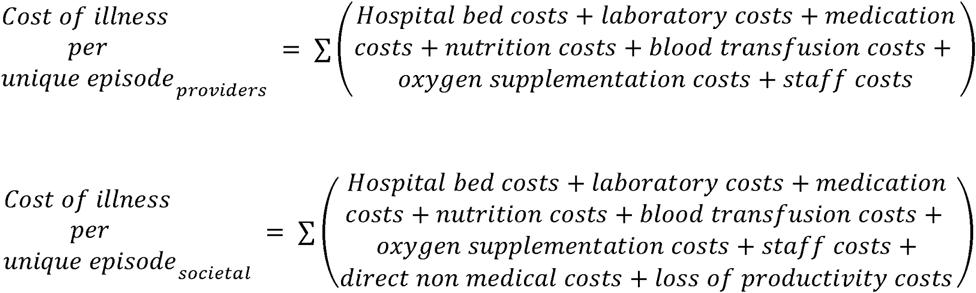

### Subgroup analysis

We estimated the mean treatment cost and mean cost for each cost category using a generalized linear model (GLM) with a gamma distribution and log-link function. This model type is ideal for right-skewed cost data [37]. The model outputs included the mean cost with 95% confidence intervals (CIs).

A subgroup analyses explored cost variations across patient characteristics, including gender, age group, geographical setting, referral status, diagnosis, and county. Appendix 5 provides a detailed justification for the selection of the reference groups. The GLM was used to estimate mean costs for each group, with inter-group cost differences assessed for statistical significance (p<0.05). Median costs were also reported and compared across subgroups using the Wilcoxon rank-sum test, with p-values used to determine if the differences were statistically significant. All analyses were performed using STATA v17 [21].

### Sensitivity analysis

To assess the robustness of our cost estimates, a one-way sensitivity analysis was conducted, from both healthcare provider and societal perspectives. The four selected parameters were varied individually while holding all other variables constant. Hospital bed day costs and staff costs were selected as they were the key drivers for the cost of treatment, whereas direct non-medical and loss of productivity costs were varied to represent literature-based assumptions.

For hospital bed day costs, the 25th (lower limit) and 75th (upper limit) percentile values were used, as described in Appendix 1. For staff costs, reported expert time estimates were adjusted using a factor derived from literature [38], as described in Appendix 6.

Direct non-medical costs were varied using their lower and upper confidence-interval values, whereas productivity-loss costs were increased by 20 percent from the base value to obtain the upper limit and reduced by 20 percent to obtain the lower limit.

### Catastrophic health expenditure

We estimated the incidence of CHE among typhoid patients using a simulation-based approach. A modified version of the methodology outlined by Xu et al. [39] was used, where the share of the total cost (direct medical and direct non-medical costs) a patient’s household would have paid OOP was varied across six different OOP share values: 20%, 24.2%, 40%, 60%, 80%, and 100%. The 24.2% share value was used as the base case, as it is the reported average OOP health expenditure in Kenya [40]. The NFHE for each wealth quintile for households from urban and rural settings was calculated using Kenyan data from national surveys [41–45]. This approach was used as primary data on the amount paid OOP by a patient’s household or the non-food household expenditure (NFHE) was not available. The detailed methodology is provided in Appendix 7.

A patient’s household experienced CHE, if the CHE ratio (*R*) of OOP to NFHE was greater than 0.4 [39]. The *R* was computed as:

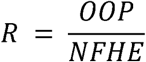

Each typhoid patient’s household was simulated across the thirty unique scenarios (six different OOP shares by five different NFHE quintiles) to determine under which scenario a patient’s household would experience CHE. A patient’s household was assigned 1 if it experienced CHE, otherwise 0. The households experiencing CHE were then summed across each unique scenario and results were disaggregated by geographical setting.

To the best of our knowledge, no published study has used the same simulation framework that we have proposed, where both the proportion of OOP payment share and the NFHE quintile were jointly varied across all possible unique scenarios. A study by Lukwa et al. used a similar approach to calculate quintile level expenditure using national survey data, but differed from our methodology as it varied the CHE threshold instead of the OOP share [46].

### Patient and public involvement

Patients were not directly involved in the design, conduct, or dissemination of this study. The study used anonymised patient-level data extracted from the CIN database. As the research relied on retrospectively collected hospital data, direct engagement with patients was not feasible. Expert opinion was incorporated through interviews with healthcare workers. All experts gave verbal consent before being interviewed.

## Results

### Patient demographics

A total of 157 children diagnosed with typhoid fever from 16 different public hospitals were included in the analysis (see Appendix 8). The median age was 8 years [IQR: 5-10 years], with just over half the patients were male (54.1%). A majority of the patients resided in rural areas (78.3%), with Busia accounting for the largest share of patients (24.2%). The median number of days a patient was hospitalized for was 3 days [IQR: 2-4 days] (Mean: 3.6 days (Range: 1-17days)).

### Cost of treatment

The median total cost of treatment from the healthcare provider’s perspective was USD 79.68 (mean: USD 95.43) per unique episode. Staff costs accounted for the largest proportion making up about 65-67% of the overall cost, with hospital bed costs (18%) being another significant cost driver. The costs for blood transfusions, oxygen supplementation, and nutritional items were grouped as “other costs” with a reported mean of USD 0.53 (mean) and median of USD 0.00 (as services were not routinely utilized). This is further detailed in Appendix 9.

From the societal perspective, the median cost of treatment was 22% higher (USD 96.95). Staff cost and hospital bed costs were still the highest contributors to the overall costs. Productivity loss and direct non-medical costs accounted for 12% and 6.7% respectively. This is reported in Table 1.

**Table 1:** Median cost of treatment from the societal perspective.

|  | GLM Mean |  |  | Median |  |  |
| --- | --- | --- | --- | --- | --- | --- |
|  | Cost (CI)<br>KES | Cost (CI)<br>USD | Share (%) | Cost [IQR]<br>KES | Cost [IQR]<br>USD | Share (%) |
| Hospital bed cost | **2,225.32**<br>(1,981.70 - 2,468.93) | **17.20**<br>(15.31 - 19.08) | 14.2 | **1,875.00**<br>[1,250.00, 2,500.00] | **14.49**<br>[9.66, 19.32] | 14.9 |
| Staff cost | **8,109.60**<br>(7,257.04 - 8,962.15) | **62.73**<br>(56.13 - 69.32) | 51.8 | **6,883.62**<br>[4,696.37, 9,070.87] | **53.25**<br>[36.33, 70.16] | 54.9 |
| Laboratory cost | **1,097.96**<br>(945.24 - 1,250.69) | **8.49**<br>(7.31, 9.67) | 7 | **914.27**<br>[554.10, 1,025.09] | **7.07**<br>[4.29, 7.93] | 7.3 |
| Medication cost | **836.13**<br>(689.78 - 982.48) | **6.47**<br>(5.34 - 7.60) | 5.3 | **564.60**<br>[253.75, 1,022.70] | **4.37**<br>[1.96, 7.91] | 4.5 |
| Other cost* | **68.70**<br>(21.19 - 116.22) | **0.53**<br>(0.16 - 0.90) | 0.4 | **0.00**<br>[0.00, 0.00] | **0.00**<br>[0.00, 0.00] | 0 |
| Direct non-medical cost | **1277.32**<br>(1096.97 - 1457.68) | **9.88**<br>(8.49 - 11.28) | 8.2 | **845.49**<br>[563.66, 1409.15] | **6.54**<br>[4.36, 10.90] | 6.7 |
| Productivity loss | **2,046.72**<br>(1,787.62 - 2,305.82) | **15.83**<br>(13.83 - 17.84) | 13.1 | **1,501.12**<br>[917.48, 2,293.70] | **11.61**<br>[4.72, 11.79] | 12.0 |
| Total cost per unique episode | **15661.75**<br>(14075.14 - 17248.35) | **121.15**<br>(108.87 - 133.42) | 100 | **12533.67**<br>[8850.90, 18472.27] | **96.95**<br>[68.46, 142.89] | 100** |

\* Other costs included blood transfusion, oxygen supplementation, and nutrition items
\*\* Median cost shares may not sum to exactly 100% due to rounding and independent calculation of component medians.

### Subgroup analysis

Table 2 summarizes the variation in the cost of typhoid treatment from the societal perspective across the different subgroups. The results from the healthcare provider perspective are presented in Appendi× 10.

**Table 2:**
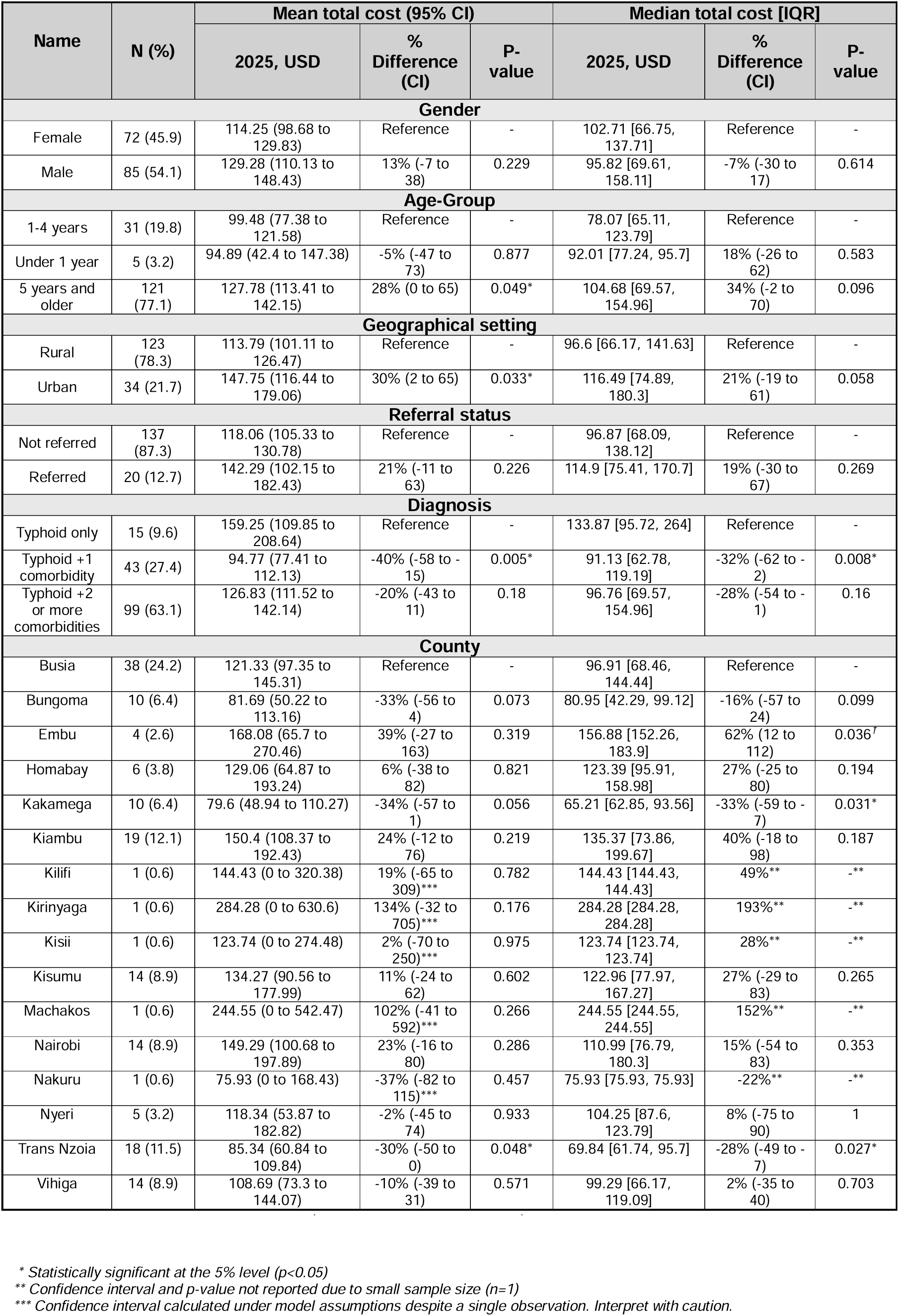
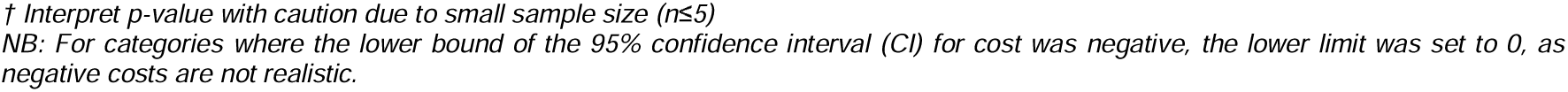
Sub-group analysis (societal perspective)

| Name | N (%) | Mean total cost (95% CI) |  |  | Median total cost [IQR] |  |  |
| --- | --- | --- | --- | --- | --- | --- | --- |
|  |  | 2025, USD | % Difference (CI) | P-value | 2025, USD | % Difference (CI) | P-value |
| Gender |  |  |  |  |  |  |  |
| Female | 72 (45.9) | 114.25 (98.68 to 129.83) | Reference | - | 102.71 [66.75, 137.71] | Reference | - |
| Male | 85 (54.1) | 129.28 (110.13 to 148.43) | 13% (-7 to 38) | 0.229 | 95.82 [69.61, 158.11] | -7% (-30 to 17) | 0.614 |
| Age-Group |  |  |  |  |  |  |  |
| 1-4 years | 31 (19.8) | 99.48 (77.38 to 121.58) | Reference | - | 78.07 [65.11, 123.79] | Reference | - |
| Under 1 year | 5 (3.2) | 94.89 (42.4 to 147.38) | -5% (-47 to 73) | 0.877 | 92.01 [77.24, 95.7] | 18% (-26 to 62) | 0.583 |
| 5 years and older | 121 (77.1) | 127.78 (113.41 to 142.15) | 28% (0 to 65) | 0.049* | 104.68 [69.57, 154.96] | 34% (-2 to 70) | 0.096 |
| Geographical setting |  |  |  |  |  |  |  |
| Rural | 123 (78.3) | 113.79 (101.11 to 126.47) | Reference | - | 96.6 [66.17, 141.63] | Reference | - |
| Urban | 34 (21.7) | 147.75 (116.44 to 179.06) | 30% (2 to 65) | 0.033* | 116.49 [74.89, 180.3] | 21% (-19 to 61) | 0.058 |
| Referral status |  |  |  |  |  |  |  |
| Not referred | 137 (87.3) | 118.06 (105.33 to 130.78) | Reference | - | 96.87 [68.09, 138.12] | Reference | - |
| Referred | 20 (12.7) | 142.29 (102.15 to 182.43) | 21% (-11 to 63) | 0.226 | 114.9 [75.41, 170.7] | 19% (-30 to 67) | 0.269 |
| Diagnosis |  |  |  |  |  |  |  |
| Typhoid only | 15 (9.6) | 159.25 (109.85 to 208.64) | Reference | - | 133.87 [95.72, 264] | Reference | - |
| Typhoid +1 comorbidity | 43 (27.4) | 94.77 (77.41 to 112.13) | -40% (-58 to -15) | 0.005* | 91.13 [62.78, 119.19] | -32% (-62 to -2) | 0.008* |
| Typhoid +2 or more comorbidities | 99 (63.1) | 126.83 (111.52 to 142.14) | -20% (-43 to 11) | 0.18 | 96.76 [69.57, 154.96] | -28% (-54 to -1) | 0.16 |
| County |  |  |  |  |  |  |  |
| Busia | 38 (24.2) | 121.33 (97.35 to 145.31) | Reference | - | 96.91 [68.46, 144.44] | Reference | - |
| Bungoma | 10 (6.4) | 81.69 (50.22 to 113.16) | -33% (-56 to 4) | 0.073 | 80.95 [42.29, 99.12] | -16% (-57 to 24) | 0.099 |
| Embu | 4 (2.6) | 168.08 (65.7 to 270.46) | 39% (-27 to 163) | 0.319 | 156.88 [152.26, 183.9] | 62% (12 to 112) | 0.036<sup>f</sup> |
| Homabay | 6 (3.8) | 129.06 (64.87 to 193.24) | 6% (-38 to 82) | 0.821 | 123.39 [95.91, 158.98] | 27% (-25 to 80) | 0.194 |
| Kakamega | 10 (6.4) | 79.6 (48.94 to 110.27) | -34% (-57 to 1) | 0.056 | 65.21 [62.85, 93.56] | -33% (-59 to -7) | 0.031* |
| Kiambu | 19 (12.1) | 150.4 (108.37 to 192.43) | 24% (-12 to 76) | 0.219 | 135.37 [73.86, 199.67] | 40% (-18 to 98) | 0.187 |
| Kilifi | 1 (0.6) | 144.43 (0 to 320.38) | 19% (-65 to 309)*** | 0.782 | 144.43 [144.43, 144.43] | 49%** | -** |
| Kirinyaga | 1 (0.6) | 284.28 (0 to 630.6) | 134% (-32 to 705)*** | 0.176 | 284.28 [284.28, 284.28] | 193%** | -** |
| Kisii | 1 (0.6) | 123.74 (0 to 274.48) | 2% (-70 to 250)*** | 0.975 | 123.74 [123.74, 123.74] | 28%** | -** |
| Kisumu | 14 (8.9) | 134.27 (90.56 to 177.99) | 11% (-24 to 62) | 0.602 | 122.96 [77.97, 167.27] | 27% (-29 to 83) | 0.265 |
| Machakos | 1 (0.6) | 244.55 (0 to 542.47) | 102% (-41 to 592)*** | 0.266 | 244.55 [244.55, 244.55] | 152%** | -** |
| Nairobi | 14 (8.9) | 149.29 (100.68 to 197.89) | 23% (-16 to 80) | 0.286 | 110.99 [76.79, 180.3] | 15% (-54 to 83) | 0.353 |
| Nakuru | 1 (0.6) | 75.93 (0 to 168.43) | -37% (-82 to 115)*** | 0.457 | 75.93 [75.93, 75.93] | -22%** | -** |
| Nyeri | 5 (3.2) | 118.34 (53.87 to 182.82) | -2% (-45 to 74) | 0.933 | 104.25 [87.6, 123.79] | 8% (-75 to 90) | 1 |
| Trans Nzoia | 18 (11.5) | 85.34 (60.84 to 109.84) | -30% (-50 to 0) | 0.048* | 69.84 [61.74, 95.7] | -28% (-49 to -7) | 0.027* |
| Vihiga | 14 (8.9) | 108.69 (73.3 to 144.07) | -10% (-39 to 31) | 0.571 | 99.29 [66.17, 119.09] | 2% (-35 to 40) | 0.703 |

\* Statistically significant at the 5% level ( $p < 0.05$ )
\*\* Confidence interval and p-value not reported due to small sample size ( $n=1$ )
\*\*\* Confidence interval calculated under model assumptions despite a single observation. Interpret with caution.
† Interpret p-value with caution due to small sample size ( $n \leq 5$ )
NB: For categories where the lower bound of the 95% confidence interval (CI) for cost was negative, the lower limit was set to 0, as negative costs are not realistic.

The cost difference between males and females was not significant. In terms of age, children older than 5 years had a 28% higher mean cost of treatment than the reference group (1-4 years), with the difference being statically significant. Urban patients incurred significantly higher median costs than those from rural areas. Referred patients had a 19% higher median cost of treatment when compared to direct admissions. Diagnosis type had a pronounced effect, with patients diagnosed with typhoid and one other comorbidity having 40% lower mean cost (statistically significant) when compared to the reference group of typhoid only diagnosis. Patients from Trans Nzoia (-28% p=0.027) and Kakamega (−33%, p=0.031) had lower median costs compared to Busia (reference), while Embu (62%, p=0.036) was the only county with a statistically significant higher median cost.

### Sensitivity analysis

Adjusting the staff time increased the median costs to USD□150.29 from the healthcare providers perspective and to USD□173.97 from the societal perspective.

When hospital bedl⍰day costs were set to the lower limit, the healthcare provider costs reduced marginally to USD□72.43 and societal costs to USD 95.89. For the upper limit, the median healthcare provider costs increased to USD 83.74 and societal costs to USD 107.09. The findings were not very sensitive to the two cost parameters obtained from literature. The detailed results are presented in Appendix□9 and illustrated in Figure 2.

**Figure 2:**
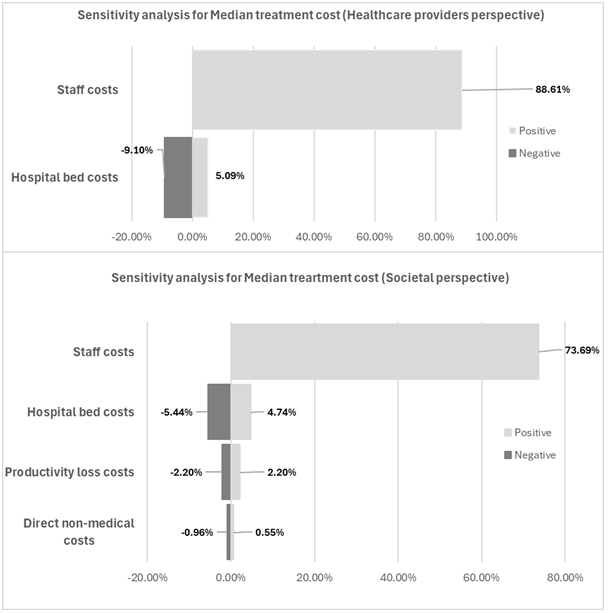
Sensitivity analysis of median cost per typhoid patient from a healthcare providers and societal perspective across parameter variations

### Catastrophic Health Expenditure

CHE occurrence increased based on the share of treatment cost paid OOP, geographical setting, and household wealth quintile. Up to an OOP share of 24.2%, no households experienced CHE. However, as the OOP share increased further, the incidence of CHE rose. At higher OOP shares (≥60%), the burden of CHE was more widespread, with rural households across the first three quintiles and urban households in the lowest quintile experiencing CHE. If households were required to pay the full cost of treatment out of pocket, rural households belonging to Q1-Q4 would experience CHE. In contrast, urban households belonging to Q2-Q5 would not face CHE irrespective of the share of OOP they had to incur. These findings are illustrated in Figure S1 in Appendix 12, with detailed estimates provided in Table 3.

**Table 3:** Patients Experiencing CHE by OOP Share and Household Wealth Quintile.

|  | Quintile | Total number of patients | OOP share (20%) | OOP share (24.2%) | OOP share (40%) | OOP share (60%) | OOP share (80%) | OOP share (100%) |
| --- | --- | --- | --- | --- | --- | --- | --- | --- |
| **Rural** | Poorest (Q1) | 123 | 0 (0%) | 0 (0%) | 1 (0.8%) | 9 (7.3%) | 14 (12.2%) | 29 (23.6%) |
|  | Poorer (Q2) |  | 0 (0%) | 0 (0%) | 0 (0%) | 1 (0.8%) | 8 (7.3%) | 13 (10.6%) |
|  | Middle (Q3) |  | 0 (0%) | 0 (0%) | 0 (0%) | 1 (0.8%) | 2 (1.6%) | 8 (6.5%) |
|  | Richer (Q4) |  | 0 (0%) | 0 (0%) | 0 (0%) | 0 (0%) | 1 (0.8%) | 1 (.8%) |
|  | Richest (Q5) |  | 0 (0%) | 0 (0%) | 0 (0%) | 0 (0%) | 0 (0%) | 0 (0%) |
| **Urban** | Poorest (Q1) | 34 | 0 (0%) | 0 (0%) | 0 (0%) | 4 (11.8%) | 7 (20.6%) | 9 (26.5%) |
|  | Poorer (Q2) |  | 0 (0%) | 0 (0%) | 0 (0%) | 0 (0%) | 0 (0%) | 0 (0%) |
|  | Middle (Q3) |  | 0 (0%) | 0 (0%) | 0 (0%) | 0 (0%) | 0 (0%) | 0 (0%) |
|  | Richer (Q4) |  | 0 (0%) | 0 (0%) | 0 (0%) | 0 (0%) | 0 (0%) | 0 (0%) |
|  | Richest (Q5) |  | 0 (0%) | 0 (0%) | 0 (0%) | 0 (0%) | 0 (0%) | 0 (0%) |

## Discussion

We estimated the cost of typhoid illness among hospitalized children in Kenya using patient-level data from the CIN, from 16 public hospitals across 16 counties in Kenya. By including both the healthcare provider and societal perspective, the analysis provided a comprehensive view of the economic burden of typhoid. The median cost of treatment from the healthcare providers’ perspective was USD 79.68 (Mean: USD 95.43) and USD 96.95 (Mean: USD 121.15) from the societal perspective.

Our estimates were consistent with those reported in India (USD 98.1) and Nepal (USD 101.84) when converted to 2025 USD [47,48], but varied from estimates reported in sub-Saharan Africa [13,49]. In Malawi, treatment costs from the provider’s perspective were more than twice our estimate, likely reflecting country-specific unit costs, disease severity, and a smaller sample size [13]. The study conducted in Tanzania reported total direct costs that were less than half of ours, but total societal costs were nearly twice as high due to the inclusion of productivity losses pre- and post-hospitalization [49].

The two main cost drivers identified in our analysis were staff time and hospital bed charges. A study looking at health expenditure associated with hospitalized children in Kenya had similar findings, reporting bed charges to be the main cost driver [34]. Inefficiencies in patient flow or delays in clinical decision-making, which are common in LMICs, can increase hospital stays, which in turn would inflate these costs [50,51]. Staff time was the highest cost driver, reflecting the labour-intensive nature of the disease, the need for a multidisciplinary approach to care, and higher salaries linked to skilled healthcare workers [52,53].

The subgroup analysis highlighted important cost variations. Children aged over 5 years and those from urban areas had significantly higher costs due to longer duration of hospitalization and greater direct non-medical and loss of productivity costs. Patients with typhoid as the only diagnosis incurred the highest cost, suggesting that a small number of severe typhoid cases drove up the costs. This is supported by the wide interquartile range in the typhoid-only subgroup. There was a significant inter-county difference in the cost of treatment from the societal perspective. Higher drug and direct non-medical costs in Embu contributed to the elevated overall costs, whereas Kakamega and Trans Nzoia had consistently lower costs across three categories (drugs, laboratory, and direct non-medical costs), suggesting less severe cases.

The sensitivity analysis underscored the importance of the staffing assumptions made in our analysis. Adjusting staff time to account for understaffing increased the total cost by nearly 90%. Kenya has only reached 68% of the Sustainable Development Goals health workforce threshold as per WHO [54]. This highlights the true economic burden of typhoid when optimal staffing care can be provided. In contrast, adjusting hospital bed-day costs had a smaller effect.

With the reported average OOP health expenditure in Kenya at 24.2% [40], our findings suggest that no household would experience CHE at this level. However, households could experience CHE, once OOP payments share reached 40%. If households were required to pay 100% of the treatment cost out of pocket, then up to 24% of rural households and about 27% of urban households could face CHE. These results highlight the need for policymakers to strengthen financial protection mechanisms through increased health insurance coverage and subsidies. While CHE incidence at the national OOP average appears reassuring, it may mask the higher risk faced by poorer households, especially those from the rural setting.

This study had some limitations. The dataset only covered the period a patient was hospitalized for, potentially underestimating the economic burden as pre- and post-hospitalization costs were not included. All patients with a diagnosis of typhoid were assumed to have been confirmed through a blood culture, in line with national clinical guidelines [55]. Finally, simulating out-of-pocket payment shares across wealth quintiles relied on hypothetical scenarios and proxy data, which may not have fully captured the real-world variations in actual healthcare costs paid OOP and household expenditure patterns.

Despite these limitations, the study has notable strengths. To our knowledge, it is the first typhoid cost-of-illness study done in Kenya and included data from 157 inpatients, which was a much larger sample than similar studies in sub-Saharan Africa [13,49]. By including patients from both urban and rural settings, the analysis captured different healthcare contexts and provides sub-national level evidence for decision-makers. The use of patient-level data and local prices for medications, diagnostics, bed-day charges, and other items provides a context-specific estimate. Calculating treatment costs from both the provider and societal perspectives gave a more comprehensive picture of typhoid’s economic burden. Finally, using a unique modeling methodology by simulating out-of-pocket payments across varying wealth levels provides policymakers with evidence on financial vulnerability. It also offers a replicable approach that other researchers can adopt in future studies where data on household expenditure and OOP for healthcare is unavailable.

Future studies should include pre- and post-admission costs, as well as the cost of care at private facilities, to better reflect the full range of typhoid-related costs. A prospective study design would allow for more accurate estimates of OOP and household expenditures, resulting in a more precise CHE calculation. Expanding the scope to include, drug-resistant, adult and outpatient cases would provide a broader picture of typhoid’s economic burden across age groups and healthcare settings.

## Conclusion

These findings provide important evidence on the costs and cost drivers of typhoid management, helping to inform future research and policy on strategies to reduce the economic burden of typhoid in Kenya. It also provides policymakers with an understanding of CHE because the disease would be experienced by households belonging to different wealth quintiles. The comprehensive, context-specific cost-of-illness estimates can be used as inputs in the economic model for the cost-effectiveness analysis of interventions such as the typhoid conjugate vaccine.

## Data Availability

All data produced in the present study are available upon reasonable request to the authors.

## Contributions

EB, PM, SO, FM, OO and LC conceptualized the study. PM led the data analysis and was supported by SO, IW, and FO. PM drafted the initial manuscript which was subsequently revised for important intellectual content by all authors. All authors read and approved the final manuscript. The work was supervised by FO. FM, OO, LC, SO, and EB. While EB, SO, FM, and OO obtained the funding for this work. PM is responsible for the overall content as guarantor.

## Ethics approval

Ethical approval was obtained from the KEMRI Scientific and Ethical Review Committee, reference number KEMRI/SERU/CGMR-C/320/4983.

