## Supplementary_Materials for "Economic Burden of Typhoid among Hospitalized Children in Kenya"

**Appendix 1: Hospital bed costs for public hospitals in Kenya.**

Table S1: Unit costs for hospital bed charges in Kenyan public hospitals

| **Hospital Name** | **Hospital level** | **Daily Bed Charges**  **(KES)** | **Daily Bed Charges**  **(USD)** |
| --- | --- | --- | --- |
| Mbagathi County Hospital | Level 5 | 1000 | 7.74 |
| Naivasha Level 5 Hospital | Level 5 | 650 | 5.03 |
| Busia County Referral Hospital | Level 5 | 200 | 1.55 |
| Kisumu County Hospital | Level 5 | 750 | 5.80 |
| Nakuru Level 5 Hospital | Level 5 | 500 | 3.87 |
| Jaramogi Oginga Odinga Teaching and Referral Hospital | Level 5 | 1000 | 7.74 |
| Kiambu Level 5 Hospital | Level 5 | 800 | 6.19 |
| Machakos Level 5 Hospital | Level 5 | 200 | 1.55 |
| Kakamega County General Teaching and Referral Hospital | Level 5 | 600 | 4.64 |
| Nyeri County Referral Hospital | Level 5 | 1900 | 14.70 |
| Embu Level 5 Teaching and Referral Hospital | Level 5 | 100 | 0.77 |
| Bungoma County Referral Hospital | Level 5 | 550 | 4.25 |
| Thika Level 5 Hospital | Level 5 | 250 | 1.93 |
| Machakos Level 5 Hospital | Level 5 | 800 | 6.19 |

| **Median cost (KES)** | 625 [IQR: 312.50 - 800] |
| --- | --- |
| **Median cost (USD)** | 4.83 [IQR: 2.42 - 6.19] |

**Appendix 2: Laboratory and radiology test unit costs in Kenya**

Table S2: Unit costs for laboratory and radiology tests

| **Test name** | **Hospital 1** | **Hospital 2** | **Hospital 3** | **Hospital 4** | **Hospital 5** | **Hospital 6** | **Hospital 7** | **Others** | **Median Unit Cost**  **(KES, 2025)** | **Median Unit Cost**  **(USD, 2025)** | **Source** |
| --- | --- | --- | --- | --- | --- | --- | --- | --- | --- | --- | --- |
| **Malaria test** |  |  |  |  |  |  |  |  |  |  |  |
| Blood slide | 100.00 | 100.00 | 50.00 | 100.00 | 100.00 | 50.00 | 100.00 |  | 110.82 | 0.86 | Public health facilities in 2023 |
| Both (Blood slide + Rapid) | 200.00 | 200.00 | 100.00 | 200.00 | 200.00 | 100.00 | 200.00 |  | 221.64 | 1.71 | Public health facilities in 2023 |
| **Haematology test** |  |  |  |  |  |  |  |  |  | ` |  |
| Full Hemogram | 600.00 | 1000.00 | 500.00 | 500.00 | 400.00 | 500.00 | 500.00 |  | 554.10 | 4.29 | Public health facilities in 2023 |
| Hb | 200.00 | 200.00 | 100.00 | 100.00 | 200.00 | 150.00 | 150.00 |  | 174.15 | 1.35 | Public health facilities in 2023 |
| **RBS** |  |  |  |  |  |  |  |  |  |  |  |
| Strip | 150.00 | 200.00 | 150.00 | 100.00 | 100.00 | 150.00 | 150.00 |  | 166.23 | 1.29 | Public health facilities in 2023 |
| Laboratory |  |  |  |  |  | 400.00 |  |  | 443.28 | 3.43 | Public health facilities in 2023 |
| **Chemistry** |  |  |  |  |  |  |  |  |  |  |  |
| Na_K | 600.00 | 1000.00 | 400.00 | 1000.00 |  | 800.00 |  |  | 886.56 | 6.86 | Public health facilities in 2023 |
| U&C | 600.00 | - | 400.00 | 800.00 |  | 800.00 | 1000.00 |  | 886.56 | 6.86 | Public health facilities in 2023 |
| Calcium | 500.00 | 400.00 | 500.00 | 200.00 | 200.00 | 500.00 | 500.00 |  | 554.10 | 4.29 | Public health facilities in 2023 |
| Albumin | 200.00 | 300.00 | 200.00 |  |  | 400.00 |  |  | 318.61 | 2.46 | Public health facilities in 2023 |
| LFT | 1600.00 | 1000.00 | 1600.00 | 2000.00 | 1000.00 | 3000.00 | 1500.00 |  | 1773.13 | 13.72 | Public health facilities in 2023 |
| U/E/C | 1500.00 | 1600.00 | 1000.00 | 1000.00 | 200.00 | 2000.00 | 1500.00 |  | 1662.31 | 12.86 | Public health facilities in 2023 |
| **HIV** |  |  |  |  |  |  |  |  |  |  |  |
| Strip |  |  |  | 100.00 | 400.00 |  |  |  | 360.17 | 2.79 | Public health facilities in 2023 |
| PCR |  |  | 1000.00 |  |  |  |  |  | 1108.20 | 8.57 | Public health facilities in 2023 |
| **Microbiology** |  |  |  |  |  |  |  |  |  |  |  |
| Lumbar puncture | 1000.00 | 1000.00 | 500.00 | 1200.00 | 600.00 | 1000.00 |  |  | 1108.20 | 8.57 | Public health facilities in 2023 |
| Blood Culture | 2000.00 | 1500.00 | 2500.00 | 2000.00 | 1000.00 | 1000.00 | 1000.00 |  | 1741.46 | 13.47 | Public health facilities in 2023 |
| **X-Ray** |  |  |  |  |  |  |  |  |  |  |  |
| Chest | 600.00 | 600.00 | 1040.00 | 400.00 | 550.00 |  | 400.00 |  | 443.22 | 3.43 | Public health facilities in 2023 |
| Wrist | 600.00 | 600.00 |  |  |  |  |  |  | 664.92 | 5.14 |  |
| Others | 800.00 | 600.00 | 1040.00 |  |  |  |  |  | 901.34 | 6.97 | Public health facilities in 2023 |
| **Urine** |  |  |  |  |  |  |  |  |  |  |  |
| Urinalysis | 200.00 | 300.00 | 100.00 | 150.00 | 150.00 | 150.00 | 200.00 |  | 197.89 | 1.53 | Public health facilities in 2023 |
| **TB Test** |  |  |  |  |  |  |  |  |  |  |  |
| Mantoux |  |  |  |  |  |  |  | 1115.25 | 1115.25 | 8.63 | https://cerbalancetafrica.ke/media/yj4k0so1/plk-price-catalogue-2022.pdf |

*Hospital 1: Kakamega County Referral Hospital, Hospital 2: Machakos Level 5 Hospital, Hospital 3: Bungoma County Referral Hospital, Hospital 4: JOOTRH, Hospital 5: Mama Lucy K Hospital, Hospital 6: Kitale County Hospital, Hospital 7: Kisumu County Hospital, and Others: Cerba Lancet Africa*

**Appendix 3: Blood transfusion costs in Kenya**

|  | **Hospital 1 (KES)** | **Hospital 2 (KES)** | **Hospital 3 (KES)** |
| --- | --- | --- | --- |
| **Cost of laboratory tests** | 500 | 500 | 500 |
| **Cost of consumables** | 200 | 500 | 500 |
| **Total cost of blood transfusion** | 700 | 1000 | 1000 |
| **Median cost (KES)** | 1000 |  |  |
| **Median cost (USD)** | 7.74 |  |  |
| **Key** |  |  |  |
| Hospital 1 = Port Reitz Sub County Hospital | |  |  |
| Hospital 2 = Jaramogi Oginga Odinga Teaching and Referral Hospital | | | |
| Hospital 3 = Nanyuki Teaching and Referral Hospital | |  |  |

Table S3: Survey results for the unit cost of blood transfusion

**Appendix 4: Staff time spend on typhoid patient care.**

Table S4: Survey results on staff time per typhoid patient

| **Cadre** | **Unit** | **Staff 1** | **Staff 2** | **Staff 3** | **Mean** |
| --- | --- | --- | --- | --- | --- |
| Specialist time (first visit) | Minutes | 30 | 15 | 20 | 21 |
| Specialist time (subsequent visit) | Minutes | 20 | 15 | 20 | 18 |
| Clinical officer (first visit) | Minutes | 15 | 10 | 10 | 11 |
| Clinical officer (subsequent visit) | Minutes | 15 | 5 | 10 | 9 |
| Nurse time per day with patient | Minutes | 60 | 64 | 62 | 62 |
| Nutritionist | Minutes | 20 | 20 | 20 | 20 |
| Medical officer (First visit) | Minutes | 10 | 10 | 10 | 10 |
| Medical officer (subsequent visit) | Minutes | 5 | 5 | 5 | 5 |

**Appendix 5: Justification for reference category selection in subgroup analysis**

Table S5: Reference categories used in subgroup analyses and justification

| **Subgroup** | **Reference Category** | **Justification Summary** |
| --- | --- | --- |
| Gender | Female | Common practice in health studies |
| Age-group | 1-4 years | Focus on vulnerable younger children |
| Geographical setting | Rural | Standard in health economic studies done in LMIC |
| Referral status | Not referred | The larger group |
| Diagnosis | Typhoid only | Simplest cases for comparison |
| County | Busia | County with the largest sample size for stability |

**Appendix 6: Adjusted staff time estimates**

This factor was calculated by dividing the reported value for normative staff levels at Level 5 hospitals by the average staffing levels reported in the survey, thereby accounting for potential under‑ or over-staffing. Mean cadre times were multiplied by the factor to obtain the adjusted values, which were then applied in the sensitivity analysis (Appendix 6).

$$Staff Adjustment Factor = \frac{Normative Staff Level (Level 5)}{Average Reported Staff Level}$$

$$Adjusted Time =Mean Reported time per cadre \times Adjustment factor$$

Table S6: Staff time estimates after applying adjustment factors

| **Cadre of Staff** | **Base Case (minutes)** | **Average Number of healthcare workers at a level 5 hospital [KHFA]**  **(Number of staff)** | **Norms and Standards (Number of staff)** | **Adjustment Factor (Norms and Standards/Average KHFA)** | **Adjusted Value (minutes)** |
| --- | --- | --- | --- | --- | --- |
| Paediatrician (first visit) | 21 | 2.3 | 9.0 | 3.91 | 81 |
| Paediatrician (subsequent visit) | 18 | 2.3 | 9.0 | 3.91 | 71 |
| Medical officer (First visit) | 10 | 23.1 | 45.0 | 1.95 | 19 |
| Medical officer (subsequent visit) | 5 | 23.1 | 45.0 | 1.95 | 10 |
| Clinical officer (first visit) | 11 | 35.3 | 30.0 | 0.85 | 10 |
| Clinical officer (subsequent visit) | 9 | 35.3 | 30.0 | 0.85 | 8 |
| Nurse | 62 | 19.8 | 20.0 | 1.01 | 63 |
| Nutritionist | 20 | 5.9 | 5.0 | 0.85 | 17 |

**Appendix 7: Detailed Methodology for CHE Analysis**

We estimated the incidence of catastrophic health expenditure (CHE) for each patient using a simulation-based approach. Each patient’s household was simulated across 30 unique scenarios, created by varying both the share of the treatment cost that would have been paid out of pocket (OOP) and the wealth quintile the household would belong to. The household’s capacity to pay (CTP) was represented by the non-food household expenditure (NFHE) for each wealth quintile. As no patient level data was available on the amount paid OOP or the NFHE, these variables were calculated using data obtained from national surveys and published literature. All monetary values obtained from literature were first converted to 2025 KES equivalents.

Below we describe the steps followed to calculate the projected OOP expenditure for the different share percentages and the NFHE for each wealth quintile.

**1. Estimation of out-of-pocket expenditure**

To determine the total OOP amount paid by each patient (p) we first obtained the total cost for each patient:

$${TotalCost}_{p} = {DirectMedicalCost}_{p} + {DirectNonMedicalCost}_{p}$$

For the different OOP share percentages, the projected OOP expenditure was calculated as:

$${OOP}_{p,s} = {TotalCost}_{p} \times{Share}_{s}$$

where *Share_s_* values were 0.20, 0.242, 0.40, 0.60, 0.80, and 1.00. The 0.242 value represented the base case, as it is the reported average OOP health expenditure in Kenya [1].

**2. Estimation of annual non-food household expenditure**

The annual NFHE for each wealth quintile (q) was calculated as per the steps described below:

**Step 1: Mean adult equivalent (AE) expenditure**

We first obtained the mean adult equivalent (AE) expenditure and its percentage shared for each quintile for both urban and rural settings from the Kenya Poverty Report 2022 [2]. The mean AE expenditure was then distributed across the different quintiles (Q1–Q5) by multiplying the mean total AE by the reported quintile percentage shares. The value for each quintile (q) was calculated as:

$${AE}_{q} = {AE}_{mean} \times\left( {{Proportion}_{q}}/{20} \right)$$

where *AE_q_* is the mean AE expenditure for quintile *q*, and *Proportion_q_* is the quintile’s proportion of total expenditure.

**Step 2: Non-food AE (monthly)**

Non-food AE expenditure (NFAE) for each quintile was then determined by multiplying the calculated *AE_q_* by the national average non-food expenditure percentage, obtained from the Kenya Poverty Report 2022 [2]:

$${NFAE}_{q} = {AE}_{q} \times Non-Food expenditure (\%)$$

**Step 3: Weighted AE factor**

The next step involved converting the household size to adult equivalent value to better estimate expenditure. This was done using adult equivalent weights and the national proportions of the three age groups (0–4 / 5–14 / 15+) per household. Data on household proportions were obtained from the 2019 Kenya National Population and Housing Census [3], and AE weights for each of the age groups were drawn from the Kenya Poverty Report 2022 [2]. A weighted AE factor (WAF) was calculated as:

$$WAF = \left( {Weight}_{0-4}\times{Proportion}_{0-4} \right) + \left( {Weight}_{5-14}\times{Proportion}_{5-14} \right)+ \left( {Weight}_{15+}\times{Proportion}_{15+} \right)$$

**Step 4: AE per household**

The resulting WAF was multiplied with the average household size to obtain the adult equivalent household size (AE_HH_):

$${AE}_{HH} = Household size \times WAF$$

The average household size was obtained from the Kenya demographic and health survey 2022 [4].

**Step 5: Monthly and Annual non-food household expenditure**

The monthly NFHE was then calculated separately for urban and rural households across the different wealth quintiles:

$${NFHE}_{monthly, q} = {AE}_{HH} \times{NFAE}_{q}$$

And this was converted to the annual NFHE:

$${NFHE}_{annual,q} = {NFHE}_{monthly,q} \times12$$

**3. Determination of catastrophic health expenditure (CHE)**

We then calculated the CHE ratio (*R_p,s_*) for each patient (*p*) across the different unique scenarios as follows:

$$R_{p,s} = \frac{{OOP}_{p,s}}{{NFHE}_{annual,q}}$$

A household was classified to have incurred CHE if the amount paid OOP was greater than 40% of the non-food household expenditure as per Xu et al.:

*CHE has occurred if* $R_{p,s} >0.4$

The total number households experiencing CHE under each unique scenario were summed and reported separately for patients from urban and rural settings.

Table S7 lists the data sources for the parameters and Table S8 summarizes the derived NFHE by quintile and geographical setting.

Table S7: Parameter inputs used to derive non-food household expenditure

| **Parameter name** | **Rural** | **Urban** | **Notes** | **Source** |
| --- | --- | --- | --- | --- |
| **Mean AE expenditure (KES, 2025)** | 6,717 | 15,261 | Converted reported value to 2025, KES. | Kenya Poverty Report 2022 [2] |
| **Quintile shares of consumption (used to derive quintile means)**  *Poorest (Q1) to Richest (Q5)* | 8.5%, 11.4%, 14.4%, 20.4%, 45.3% | 3.7%, 11.5%, 15.9%, 25.5%, 43.4% |  | Kenya Poverty Report 2022 [2] |
| **Non-food %** | 34.2 | 55.2 |  | Kenya Poverty Report 2022 [2] |
| **Household (HH) size**  **(persons)** | 4.41 | 3.12 |  | Kenya demographic and health survey 2022 [4] |
| **National Age proportions by age group  (0–4 / 5–14 / 15+)** | 12.7 / 28.9 / 58.4 | 12.3 / 20.8 / 66.9 |  | 2019 Kenya population and housing census [3] |
| **AE weights by age-group  (0–4 / 5–14 / 15+)** | 0.24 / 0.65 / 1.0 | 0.24 / 0.65 / 1.0 |  | Kenya Poverty Report 2022 [2] |
| **Income per month  per earner (15+) (2025, KES)** | Low: 9,052 Average: 21,053 High: 33,783 | Low: 16,967 Average: 26,641 High: 38,287 | Converted reported value to 2025, KES. | Providing, managing and promoting quality statistics (Economic Survey 2024) [5] |
| **Employment to population ratio, 15+, total (%) (modeled ILO estimate) - Kenya** | 63 | 63 | 2024 national value | World Bank Open Data [6] |

Table S8: Derived non-food household expenditure by quintile and residence type (monthly and annual)

| **Residence type** | **Quintile** | **Mean AE expenditure (KES, 2025)** | **Proportion (%)** | **Total expenditure  per month per AE (KES, 2025)** | **Non-food proportion (%)** | **Non-food per AE  per month (KES, 2025)** | **HH  size  (persons)** | **Proportion per  age-group  (0–4 / 5–14 / 15+)** | **AE weights by  age-group  (0–4 / 5–14 / 15+)** | **Weighted AE**  **factor** | **AE  per  HH** | **Non-food expenditure per household per month  (KES, 2025)** | **Non-food expenditure per household per year  (KES, 2025)** |
| --- | --- | --- | --- | --- | --- | --- | --- | --- | --- | --- | --- | --- | --- |
| **Rural** | Q1 | 6717 | 8.5 | 2,855 | 34.2 | 976 | 4.41 | 12.6 / 26.3 / 61.1 | 0.24 / 0.65 / 1.0 | 0.80233 | 3.54 | 3,454 | 41,454 |
|  | Q2 |  | 11.4 | 3,829 |  | 1309 |  |  |  |  |  | 4,633 | 55,597 |
|  | Q3 |  | 14.4 | 4,836 |  | 1654 |  |  |  |  |  | 5,852 | 70,227 |
|  | Q4 |  | 20.4 | 6,851 |  | 2343 |  |  |  |  |  | 8,291 | 99,489 |
|  | Q5 |  | 45.3 | 15,214 |  | 5203 |  |  |  |  |  | 18,410 | 220,924 |
| **Urban** | Q1 | 15261 | 3.7 | 2,823 | 55.2 | 1558 | 3.12 | 12.6 / 26.3 / 61.1 | 0.24 / 0.65 / 1.0 | 0.83372 | 2.60 | 4,054 | 48,646 |
|  | Q2 |  | 11.5 | 8,775 |  | 4844 |  |  |  |  |  | 12,600 | 151,198 |
|  | Q3 |  | 15.9 | 12,132 |  | 6697 |  |  |  |  |  | 17,421 | 209,048 |
|  | Q4 |  | 25.5 | 19,458 |  | 10741 |  |  |  |  |  | 27,939 | 335,265 |
|  | Q5 |  | 43.4 | 33,116 |  | 18280 |  |  |  |  |  | 47,551 | 570,608 |

**Appendix 8: Demographic and referral characteristics of the study population**

Table S9: Demographic and referral characteristics of the study population

| **Name** | **Patient count** | **Percent (%)** |
| --- | --- | --- |
| **Gender** |  |  |
| Male | 85 | 54.1 |
| Female | 72 | 45.9 |
| **Age-group** |  |  |
| Under 1 year | 5 | 3.2 |
| 1-4 years | 31 | 19.8 |
| 5 years and older | 121 | 77.1 |
| **Setting type** |  |  |
| Rural | 123 | 78.3 |
| Urban | 34 | 21.7 |
| **Hospital county** |  |  |
| Bungoma | 10 | 6.4 |
| Busia | 38 | 24.2 |
| Embu | 4 | 2.6 |
| Homabay | 6 | 3.8 |
| Kakamega | 10 | 6.4 |
| Kiambu | 19 | 12.1 |
| Kilifi | 1 | 0.6 |
| Kirinyaga | 1 | 0.6 |
| Kisii | 1 | 0.6 |
| Kisumu | 14 | 8.9 |
| Machakos | 1 | 0.6 |
| Nairobi | 14 | 8.9 |
| Nakuru | 1 | 0.6 |
| Nyeri | 5 | 3.2 |
| Trans Nzoia | 18 | 11.5 |
| Vihiga | 14 | 8.9 |
| **Referral status** |  |  |
| Referred to the hospital | 20 | 12.7 |
| Not referred to the hospital | 137 | 87.3 |

**Appendix 9: Median cost of treatment from the healthcare provider’s perspective**

Table S10: Median cost of treatment from the healthcare provider’s perspective

|  | **GLM Mean** | | | **Median** | | |
| --- | --- | --- | --- | --- | --- | --- |
|  | **Cost** (CI)  **KES** | **Cost** (CI)  **USD** | **Share (%)** | **Cost** [IQR]  **KES** | **Cost** [IQR] **USD** | **Share (%)** |
| **Hospital bed cost** | **2,225.32**  (1,981.70 - 2,468.93) | **17.20**  (15.31 - 19.08) | 18 | **1,875.00**  [1,250.00, 2,500.00] | **14.49**  [9.66, 19.32] | 18.2 |
| **Staff cost** | **8,109.60**  (7,257.04 - 8,962.15) | **62.73**  (56.13 - 69.32) | 65.7 | **6,883.62**  [4,696.37, 9,070.87] | **53.25**  [36.33, 70.16] | 66.8 |
| **Laboratory cost** | **1,097.96**  (945.24 - 1,250.69) | **8.49**  (7.31 - 9.67) | 8.9 | **914.27**  [554.10, 1,025.09] | **7.07**  [4.29, 7.93] | 8.9 |
| **Medication cost** | **836.13**  (689.78 - 982.48) | **6.47**  (5.34 - 7.60) | 6.8 | **564.60**  [253.75, 1,022.70] | **4.37**  [1.96, 7.91] | 5.5 |
| **Other cost*** | **68.70**  (21.19 - 116.22) | **0.53**  (0.16 - 0.90) | 0.6 | **0.00**  [0.00, 0.00] | **0.00**  [0.00, 0.00] | 0.0 |
| **Total cost per unique episode** | **12,337.71**  [11,144.94, 13,530.48] | **95.43**  (86.21 - 104.66) | 100 | **10,301.24**  [7,009.54, 15,150.18] | **79.68**  [54.22, 117.19] | 100** |

** Other cost includes blood transfusion, oxygen supplementation, and nutrition items*

*** Median cost shares may not sum to exactly 100% due to rounding and independent calculation of component medians.*

**Appendix 10: Subgroup analysis of treatment costs from the healthcare provider perspective**

Table S11: Treatment cost differences across subgroups from the healthcare provider perspective

| **Name** | **N (%)** | **Mean total cost (95% CI)** | | | **Median total cost [IQR]** | | |
| --- | --- | --- | --- | --- | --- | --- | --- |
|  |  | **2025, USD** | **% Difference (CI)** | **P-value** | **2025, USD** | **% Difference (CI)** | **P-value** |
| **Gender** | | | | | | |  |
| Female | 72 (45.9) | 91.49  (79.5 to 103.48) | Reference | - | 84.69  [53.08, 103.45] | Reference | - |
| Male | 85 (54.1) | 100.09  (85.84 to 114.34) | 9%  (-10 to 33) | 0.363 | 78.52  [55.81, 126.40] | -7%  (-29% to 15%) | 0.671 |
| **Age-Group** | | | | | | | |
| 1-4 years | 31 (19.8) | 80.73  (63.48 to 97.97) | Reference | - | 62.84  [51.76, 103.19] | Reference | - |
| Under 1 year | 5 (3.2) | 74.91  (35.06 to 114.76) | -7%  (-48 to 65) | 0.798 | 74.82  [56.45, 78.51] | 19%  (-35% to 73%) | 0.599 |
| 5 years and older | 121 (77.1) | 100.05  (89.23 to 110.87) | 24%  (-2 to 57) | 0.079 | 85.52  [55.45, 123.22] | 36%  (-8% to 79%) | 0.086 |
| **Geographical setting** | | | | | | | |
| Rural | 123 (78.3) | 92.86  (82.77 to 102.96) | Reference | - | 79.42  [54.29, 117.19] | Reference | - |
| Urban | 34 (21.7) | 104.73  (83.08 to 126.39) | 13%  (-11 to 42) | 0.313 | 85.91  [53.63, 136.68] | 8%  (-24% to 41%) | 0.701 |
| **Referral status** | | | | | | | |
| Not referred | 137 (87.3) | 93.30  (83.68 to 102.91) | Reference | - | 79.68  [53.65, 106.55] | Reference | - |
| Referred | 20 (12.7) | 110.06  (80.38 to 139.75) | 18%  (-12 to 57) | 0.262 | 89.32  [56.47, 134.57] | 12%  (-39% to 63%) | 0.320 |
| **Diagnosis** | | | | | | | |
| Typhoid only | 15  (9.6) | 119.76  (84.19 to 155.32) | Reference | - | 96.07  [78.53, 180.83] | Reference | - |
| Typhoid +1 comorbidity | 43 (27.4) | 75.05  (61.88 to 88.21) | -37%  (-55 to -12) | 0.008* | 73.95  [50.28, 95.08] | -23%  (-47% to 9%) | 0.008* |
| Typhoid +2 or more comorbidities | 99 (63.1) | 100.6  (88.97 to 112.23) | -16%  (-39 to 16) | 0.284 | 79.57  [54.71, 124.79] | -17%  (-44% to 9%) | 0.213 |
| **County** | | | | | | | |
| Busia | 38 (24.2) | 99.32  (80.1 to 118.54) | Reference | - | 79.72  [57.01, 121.52] | Reference | - |
| Nakuru | 1  (0.6) | 54.12  (0 to 118.69) | -46%  (-84 to 83)*** | 0.325 | 54.12  [54.12, 54.12] | -32% | -* |
| Kakamega | 10  (6.4) | 65.85  (41.01 to 90.7) | -34%  (-57 to 1) | 0.058 | 53.76  [51.39, 76.37] | -33%  (-60% to -5%) | 0.031^†^ |
| Bungoma | 10  (6.4) | 68.51  (42.66 to 94.37) | -31%  (-55 to 5) | 0.086 | 66.93  [36.56, 84.69] | -16%  (-58% to 26%) | 0.110 |
| Embu | 4  (2.6) | 136.57  (55.1 to 218.04) | 38%  (-27 to 157) | 0.320 | 128.23  [123.62, 149.53] | 61%  (15% to 106%) | 0.036* |
| Homabay | 6  (3.8) | 106.14  (54.44 to 157.85) | 7%  (-37 to 81) | 0.804 | 100.48  [78.73, 130.33] | 26%  (-26% to 78%) | 0.194 |
| Kiambu | 19 (12.1) | 104.99  (76.25 to 133.73) | 6%  (-24 to 48) | 0.746 | 93.79  [53.07, 137.29] | 18%  (-30% to 66%) | 0.886 |
| Kilifi | 1  (0.6) | 121.52  (0 to 266.51) | 22%  (-63 to 310)*** | 0.744 | 121.52  [121.52, 121.52] | 52% | -** |
| Kirinyaga | 1  (0.6) | 238.45  (0 to 522.96) | 140%  (-28 to 704)*** | 0.156 | 238.45  [238.45, 238.45] | 199% | -** |
| Kisii | 1  (0.6) | 106.55  (0 to 233.68) | 7%  (-68 to 259)*** | 0.909 | 106.55  [106.55, 106.55] | 34% | -** |
| Kisumu | 14  (8.9) | 102.11  (69.55 to 134.67) | 3%  (-29 to 49)*** | 0.884 | 93.23  [60.98, 124.79] | 17%  (-32% to 66%) | 0.550 |
| Machakos | 1  (0.6) | 188.65  (0 to 413.74) | 90%  (-43 to 536)*** | 0.298 | 188.65  [188.65, 188.65] | 137% | -** |
| Nairobi | 14  (8.9) | 108.00  (73.56 to 142.44) | 9%  (-25 to 58)*** | 0.660 | 83.72  [54.98, 136.68] | 5%  (-51% to 62%) | 0.992 |
| Nyeri | 5  (3.2) | 100.01  (46.65 to 153.37) | 1%  (-43 to 78) | 0.981 | 92.79  [76.15, 106.60] | 16%  (-63% to 96%) | 0.880 |
| Trans Nzoia | 18 (11.5) | 71.02  (51.05 to 90.99) | -28% (-49 to 1) | 0.054 | 58.39  [50.28, 78.51] | -27%  (-48% to -5%) | 0.027* |
| Vihiga | 14  (8.9) | 89.86  (61.21 to 118.52) | -10% (-38 to 31) | 0.599 | 82.11  [54.71, 99.76] | 3%  (-34% to 40%) | 0.741 |

** Statistically significant at the 5% level (p < 0.05)*

*** Confidence interval and p-value not reported due to small sample size (n = 1)*

****Confidence interval calculated under model assumptions despite a single observation. Interpret with caution.*

*† Interpret p-value with caution due to small sample size (n ≤ 5)*

*NB: For categories where the lower bound of the 95% confidence interval (CI) for cost was negative, the lower limit was set to 0, as negative costs are not realistic.*

**Appendix 11: Sensitivity analysis of treatment costs**

Table S12: Results of sensitivity analysis (Median)

| **Medians** | | | | | | | | |
| --- | --- | --- | --- | --- | --- | --- | --- | --- |
|  | **Healthcare provider** | | | | **Societal** | | | |
|  | **KES** | **% difference** | **USD** | **% difference** | **KES** | **% difference** | **USD** | **% difference** |
| **Base case** | 10301.24 | 0.00% | 79.68 | 0.00% | 12533.67 | 0.00% | 96.95 | 0.00% |
| **Staff time (Adjusted)** | 19429.49 | 88.61% | 150.29 | 88.62% | 21769.12 | 73.69% | 168.39 | 73.69% |
| **Bed charges (Upper limit)** | 10825.91 | 5.09% | 83.74 | 5.10% | 13128.06 | 4.74% | 101.55 | 4.74% |
| **Bed charges (Lower limit)** | 10301.24 | 0.00% | 79.68 | 0.00% | 12533.67 | 0.00% | 96.95 | 0.00% |
| **Productivity loss (Lower limit)** | - | - | - | - | 12258.42 | -2.20% | 94.82 | -2.20% |
| **Productivity loss (Upper limit)** | - | - | - | - | 12808.92 | 2.20% | 99.08 | 2.20% |
| **Direct non-medical cost (Lower limit)** | - | - | - | - | 12413.43 | -0.96% | 96.02 | -0.96% |
| **Direct non-medical cost (Upper limit)** | - | - | - | - | 12603.06 | 0.55% | 97.49 | 0.55% |
| Table S13: Results of sensitivity analysis (Mean) | | | | | | | | |
| **Means** | | | | | | | | |
|  | **Healthcare provider** | | | | **Societal** | | | |
|  | **KES** | **% difference** | **USD** | **% difference** | **KES** | **% difference** | **USD** | **% difference** |
| **Base case** | 12337.71 | 0.00% | 95.43 | 0.00% | 15661.75 | 0.00% | 121.15 | 0.00% |
| **Staff time (Adjusted)** | 23157.93 | 87.70% | 179.13 | 87.71% | 26587.14 | 69.76% | 205.66 | 69.76% |
| **Bed charges (Upper limit)** | 12960.32 | 5.05% | 100.25 | 5.05% | 16389.66 | 4.65% | 126.78 | 4.65% |
| **Bed charges (Lower limit)** | 12337.71 | 0.00% | 95.43 | 0.00% | 15661.75 | 0.00% | 121.15 | 0.00% |
| **Productivity loss (Lower limit)** | - | - | - | - | 15252.40 | -2.61% | 117.98 | -2.61% |
| **Productivity loss (Upper limit)** | - | - | - | - | 16071.09 | 2.61% | 124.31 | 2.61% |
| **Direct non-medical cost (Lower limit)** | - | - | - | - | 15511.09 | -0.96% | 119.98 | -0.96% |
| **Direct non-medical cost (Upper limit)** | - | - | - | - | 15766.57 | 0.67% | 121.96 | 0.67% |

**Appendix 12: CHE results**

Urban households

Percentage of housholds facing CHE

| Household wealth quintile | 20% OOP | 24.2% OOP | 40% OOP | 60% OOP | 80% OOP | 100% OOP |
| --- | --- | --- | --- | --- | --- | --- |
| Q1 (Poorest) | 0 | 0 | 0 | 11.8 | 20.6 | 26.5 |
| Q2 (Poorer) | 0 | 0 | 0 | 0 | 0 | 0 |
| Q3 (Middle) | 0 | 0 | 0 | 0 | 0 | 0 |
| Q4 (Richer) | 0 | 0 | 0 | 0 | 0 | 0 |
| Q5 (Richest) | 0 | 0 | 0 | 0 | 0 | 0 |

Rural households

Percentage of housholds facing CHE

| Household wealth quintile | 20% OOP | 24.2% OOP | 40% OOP | 60% OOP | 80% OOP | 100% OOP |
| --- | --- | --- | --- | --- | --- | --- |
| Q1 (Poorest) | 0 | 0 | 0.8 | 7.3 | 12.2 | 23.6 |
| Q2 (Poorer) | 0 | 0 | 0 | 0.8 | 7.3 | 9.8 |
| Q3 (Middle) | 0 | 0 | 0 | 0.8 | 1.6 | 6.5 |
| Q4 (Richer) | 0 | 0 | 0 | 0 | 0.8 | 0.8 |
| Q5 (Richest) | 0 | 0 | 0 | 0 | 0 | 0 |

Figure S1: Percentage of rural and urban households with CHE by quintile

2	Kenya National Bureau of Statistics (KNBS). The Kenya Poverty Report 2022. Nairobi, Kenya: Kenya National Bureau of Statistics 2024.

3	*2019 Kenya population and housing census*. Nairobi: Kenya National Bureau of Statistics 2019.

4	*Kenya demographic and health survey, 2022*. Nairobi, Kenya: Kenya Nationa Bureau of Statistics 2023.

5	*Providing, managing and promoting quality statistics (Economic Survery 2024)*. Nairobi, Kenya: Kenya National Bureau of Statistics 2024.

6	Employment to population ratio, 15+, total (%) (modeled ILO estimate). World Bank Open Data. https://data.worldbank.org/indicator/SL.EMP.TOTL.SP.ZS (accessed 2 October 2025)
